# Performance of an Electroencephalography-Measuring Headband or Actigraphy Compared with Polysomnography in Older Adults with Sleep Disturbances

**DOI:** 10.1101/2025.01.25.25321124

**Authors:** Brienne Miner, Yulu Pan, Gawon Cho, Jarett Talarczyk, Anne Chen, Chase Burzynski, Lakshmi Polisetty, Margaret Doyle, Lynne Iannone, Slawomir Mejnartowicz, Richard A. Breier, Thomas M. Gill, Henry K. Yaggi, Melissa Knauert

**Affiliations:** Yale University School of Medicine, New Haven, Connecticut; VA Connecticut Healthcare System, West Haven, CT

**Author notes:** Corresponding Author: Brienne Miner, MD MHS; 333 Cedar Street, New Haven, CT, 06520, USA. [; @SleepinBeautyMD].

**Keywords:** sleep, aging, polysomnography, actigraphy, wearables

## Abstract

**Study Objectives:** In older adults, self-reported sleep measures may be inaccurate, but polysomnography (PSG) is burdensome. We assessed the performance of an electroencephalography-measuring headband (HB) or actigraphy (ACT) compared with PSG in older adults with sleep disturbances.

**Methods:** Sixty-three adults aged ≥60 years who reported symptoms of insomnia and/or daytime sleepiness ≥once/week completed a week-long, home-based protocol during which they wore the HB for seven nights, an actigraph for seven days and nights, and completed a one-night level II unattended PSG. For the current analysis, we compared total sleep time (TST) and wake after sleep onset (WASO) from all three devices on the PSG night. We calculated absolute differences and intraclass correlation coefficients (ICCs) for TST and WASO between HB and ACT, respectively, vs. PSG. We also evaluated the performance of the HB among subgroups of the poorest sleepers according to the presence of sleep apnea, insomnia, poor sleep quality, and periodic limb movements of sleep. Feasibility of the HB was assessed by measures of adherence (i.e., ability to use the HB over seven nights) and usability (i.e., ratings of items from the WEarable Acceptability Range [WEAR] scale).

**Results:** The average age was 72.8 [standard deviation 6.6] years, 63.5% were female, and 63.5% identified as non-Hispanic White. On PSG, averages for TST and WASO were 370.1 [93] and 88.9 [63] minutes, respectively. For the HB vs. PSG, mean differences and ICCs were -11.9 minutes and 0.83 [0.74, 0.89] for TST; and -15.5 minutes and 0.65 [0.48, 0.77] for WASO. For ACT vs. PSG, mean differences for TST and WASO were larger, and ICCs showed lower levels of agreement. The HB performed well among the poorest sleepers, with ICCs >0.65 for TST and WASO. On average, participants wore the HB for 6.5 [0.8] nights, and usability was rated highly.

**Conclusions:** The HB demonstrated good agreement with PSG, outperforming ACT, including among the poorest sleepers. Devices like the HB might provide feasible measures of sleep that are more accurate than ACT and enhance the management of sleep health in older adults with sleep disturbances. Future research should focus on further validation of these devices in habitual sleep environments.

## INTRODUCTION

Disturbances affecting the duration or quality of sleep occur in nearly half of older adults.^1,2^ These disturbances are associated with cognitive decline, depression, disability, institutionalization, and high healthcare costs,^3–7^ calling for their accurate identification and treatment. Unfortunately, self-reported sleep may not accurately reflect habitual sleep duration or quality, leading to missed treatment opportunities or overtreatment with high-risk medications.^1,2,8–11^ Polysomnography (PSG), which includes electroencephalography (EEG), is the gold standard for evaluating sleep,^12^ but it is costly, burdensome, and may not reflect habitual sleep patterns.^13–15^ Actigraphy (ACT) is the reference standard for objectively evaluating sleep over multiple nights, but it relies on activity counts to estimate sleep and wake time indirectly. As a result, ACT overestimates sleep time, underestimates time awake, and is less accurate in older adults with sleep disturbances or multiple chronic conditions.^10,11,16^ Thus, tools that feasibly and accurately characterize habitual sleep duration and quality in older adults are needed to improve the targeting and monitoring of interventions to improve sleep health and prevent adverse outcomes.

Recently, new wearable devices with EEG capability have been introduced. One such device is a multi-sensor EEG headband (HB). The HB was validated against PSG in small samples of healthy middle-aged adults (N=25 adults; average age 35 years)^17^ and older adults with Parkinson’s disease (N=10; average age 70 years),^18^ where it showed over 80% accuracy in both cases for sleep-staging compared to PSG. However, these studies excluded persons using hypnotic medications or with known insomnia or sleep apnea, resulting in diminished generalizability.^17,18^ The validity and usability of the HB in a more generalizable population of older adults with sleep deficiency needs to be demonstrated, as this group has the highest prevalence of sleep disturbances and is also likely to experience disagreement between gold-standard and actigraphy measures of sleep.^16^

In the current study, we compared the agreement of the HB vs. PSG and ACT vs. PSG, respectively, in a sample of older adults with self-reported sleep disturbances. Participants wore the HB for 7 nights, ACT for 7 days and nights, and completed an in-home PSG during one of the 7 nights. We hypothesized that measures of duration and quality from the HB, measured via EEG rather than activity counts, would have higher agreement with PSG than ACT. We also assessed the feasibility of using the HB in this population. Due to its ergonomic design, we hypothesized that participants would have high rates of adherence to the 7-night protocol and report high rates of usability.

## METHODS

### Study Population and Protocol

This is a cross-sectional study of community-living older adults in the greater New Haven area of Connecticut. Participants were ≥ 60 years old and reported sleep disturbances, including insomnia symptoms (i.e., difficulties with sleep onset, sleep maintenance, or early morning awakenings) and/or daytime sleepiness (i.e., frequent dozing, needing to nap, or feeling like one could fall asleep easily during the day) at least once a week. Participants were recruited via flyers at Yale-New Haven Hospital-affiliated primary care clinics and local senior centers. We excluded participants with severe cognitive impairment (defined as ≥4 errors on the 6-item Callahan screener),^19^ and those who could not converse in English, refused or were unable to consent, lived in an assisted living or extended care facility, did not live within driving distance, or had an unplanned hospitalization in the past month. Importantly, we did not exclude otherwise eligible persons who were taking hypnotic medications or had a known diagnosis of sleep apnea, insomnia, or restless legs syndrome. Of the 78 persons screened, 71 ultimately enrolled in the study (5 dropped before the baseline interview; 2 were not eligible [1 had an unplanned overnight hospitalization in the past month; 1 did not meet age requirements]). Among the 71 enrolled in the study, 8 did not complete the study protocol (6 were unable to use the HB; 1 had no PSG data; 1 had no ACT data). Hence, the final analytical sample included 63 participants (88.7% of persons enrolled). The Yale University Human Investigation Committee approved the study protocol, and all participants provided written informed consent.

We conducted a week-long protocol with collection of demographic and clinical information, and subjective and objective sleep measures. During the initial visit (day 1), we collected clinical information (*described below*) and introduced participants to the sleep devices (HB, ACT, PSG) to be used during the protocol. On the night of the participant’s choosing (but after several nights of accommodation to the HB), we visited participant’s homes to set up an overnight PSG. This night, the three sleep devices were worn simultaneously. The following morning, participants removed the PSG equipment and continued to wear the HB (at night) and ACT (all day) for the remainder of the protocol. Participants completed daily sleep diaries for seven days and completed a usability questionnaire for the HB at the end of the protocol, when all study devices and diaries were collected.

### Demographic and Clinical Characteristics

Demographic characteristics included age, sex, race/ethnicity (non-Hispanic White vs. other), education (college graduate vs. other), and occupational status (retired vs. other). Clinical characteristics included body mass index (BMI), cognitive impairment (Montreal Cognitive Assessment short form score < 12),^20^ self-reported physician diagnoses of diabetes, coronary artery disease, heart failure, or chronic obstructive pulmonary disease, physician diagnosis of a psychiatric condition (including depression, anxiety, bipolar disorder, post-traumatic stress disorder, or substance use disorder), and physician diagnosis of a sleep disorder (including sleep apnea, insomnia, or restless legs syndrome).

Participants were asked about their use in the previous two weeks of prescription and over-the-counter medications. Medication information was ascertained through a review of medication bottles, medication lists, or electronic medical record. For the total number of medications used, we included prescription medications and over-the-counter use of aspirin, pain medication (e.g., acetaminophen, ibuprofen, naproxen), allergy/cold medicines (e.g., diphenhydramine, cetirizine), or antacids (e.g., tums, famotidine, proton pump inhibitors). The count did not include bowel regimens, eye drops, topical lotions, nasal sprays, multivitamins or probiotics. We also inquired specifically about sleep medications by asking, “In the past two weeks, have you taken a sleep aide to help you sleep during the night? These can include a prescription or over-the-counter medication or an herbal or nutritional supplement.”

### Measures of Self-Reported and Objective Sleep

Self-reported sleep measures included the Pittsburgh Sleep Quality Index (PSQI),^21^ the Epworth Sleepiness Scale (ESS),^22^ and the Insomnia Severity Index (ISI).^23^ The PSQI assesses sleep quality over the past month (score range=0-21), with higher scores indicating worse sleep quality and scores >5 denoting poor sleep quality.^21^ The ESS measures the likelihood of dozing or sleeping in eight hypothetical situations during the day (score range= 0–24), with higher scores indicating more severe daytime sleepiness.^22^ The ISI captures symptoms and daytime consequences of insomnia based on the Diagnostic and Statistical Manual of Mental Disorders, 5th Edition, Text Revision (DSM-5-TR).^23^ ISI scores range from 0–28, with higher scores indicating more severe insomnia.^23^ An ISI score of ≥8 establishes sub-threshold insomnia, while scores ≥15 indicate moderate to severe insomnia.^23^

Objective sleep measures included the HB, ACT, and PSG. The HB (formerly from Dreem, Paris, France; now from Beacon Biosignals, Boston, USA) is a wireless, self-administered, non-invasive, FDA class II medical device. The HB measures physiological signals via five dry EEG electrodes (*described below*), a 3-dimensional accelerometer (to gauge movements, position, and breathing), and an infrared pulse oximeter.^17^ It records, stores, and automatically analyzes data in real time without any connection. The HB connects via Bluetooth to a mobile device application on a smartphone or tablet and transfers data via Wi-Fi to the sponsor’s servers. The HB electrodes are located over the anterior forehead and the back of the head, where flexible silicone protrusions enable signal acquisition from the scalp. Signal acquisition is at 250 Hz with a 0.4–35 Hz bandpass filter. The HB has five electrodes: F7 and F8 (located over the left and right frontal areas); FpZ (ground sensor located on the frontal band); and O1 and O2 (located in the left and right occipital areas). The five electrodes yield seven derivations (FpZ-O1, FpZ-O2, FpZ-F7, F8-F7, F7-O1, F8-O2, FpZ-F8). For this study, participants were not required to use a mobile device to manage the HB. During the baseline visit, participants were instructed on the placement and operation of the HB, including how to start and stop recordings manually. All data from the HB were retrieved at the end of the week-long protocol once the study devices had been collected. Participants were instructed to wear the HB for up to 7 nights, averaging 6.5 (0.8) days. For the current study, we measured total sleep time (TST; total time in minutes that a participant remained asleep at night between sleep onset and final awakening) and wake after sleep onset (WASO; amount of time that a participant was awake between sleep onset and final awakening) on the night of PSG.

We used the Actiwatch Spectrum Plus (ACT; Phillips Respironics, Murrysville, PA, U.S.A.), a standard wrist-worn actigraph. ACT estimates sleep objectively and non-invasively by integrating the occurrence and degree of limb movements.^24^ For the current study, participants were instructed to wear the ACT for 7 days and nights on the non-dominant wrist, averaging 7.2 (0.5) days worn. ACT data were analyzed using Phillips Respironics software with proportional integration mode. The software algorithm and sleep diaries were used to edit the raw data and generate different sleep metrics, including TST and WASO. For the current study, we used ACT to obtain TST and WASO on the night of PSG.

We performed a one-night, home-based PSG with the NOX-Self applied system (Nox Medical, Reykjavik, Iceland).^25^ Trained research assistants traveled to participants’ homes to set up the entirety of the PSG system for participants around their habitual sleep time. The NOX-Self Applied system provides full-montage PSG, including channels for EEG, electrooculography, electromyography, airflow sensors, chest and abdominal respiratory effort bands, EKG, and oxygen saturation. For the current study, the HB was placed over the head after PSG equipment was placed to ensure that the sensors would not interfere with each other. PSGs were scored by a trained sleep technologist at Sleep Strategies, Inc. The Apnea-Hypopnea Index (AHI) was the average number of apneas or hypopneas with at least 3% oxygen desaturation per hour during sleep. The Periodic limbic movement index (PLMI) was defined as the average number of occurrences of leg movements per hour of sleep. For the current study, we measured TST and WASO from PSG on the night when all three sleep devices were used.

### Adherence and Usability of the HB

Adherence for the HB was assessed by calculating: 1) the mean number of nights the HB was used; 2) the percentage of nights participants used the HB for at least 4 hours (on nights used); and 3) the percentage of participants who used the HB for all seven nights of the protocol. Usability of the HB was assessed by eleven items from the WEarable Acceptability Range (WEAR) scale, which includes items on the aesthetics, ergonomics, and social desirability of wearable technologies.^26,27^ Responses were recorded on a Likert scale that ranged from 1 (strongly disagree) to 5 (strongly agree).^26,27^ Participants completed the WEAR scale at the end of the 8-day protocol. Five items with a negative valence (i.e., items 2-4, 8, 10; *shown in Table 4*) were reverse coded.

### Statistical Analysis

Demographic, clinical, and sleep characteristics were described using means and standard deviations (SD) for continuous variables and counts with frequency (%) for categorical variables. To evaluate the performance of the HB and ACT compared with PSG, we used a standardized framework for testing the validity of sleep devices developed by Menghini et al.^28^ Pearson’s correlation coefficients were used to estimate the strength and direction of TST and WASO for HB vs. PSG and ACT vs. PSG. Absolute differences in TST and WASO between HB or ACT vs. PSG were analyzed using paired t-tests (difference= polysomnography – device [headband or actigraphy]). Two-way mixed effects intraclass correlation coefficients (ICCs) were used to assess the agreement between HB vs. PSG and ACT vs. PSG for TST and WASO, treating HB, ACT, and PSG as different “raters”. ICC reliability values <0.5 indicate poor agreement, 0.5-0.75 indicate moderate agreement, and values ≥0.75 indicate good to excellenagreement.^29^ Bland-Altman plots were used to graphically show the differences in TST and WASO for HB vs. PSG and ACT vs. PSG.^30^ Bland-Altman plots display the agreement between two different measurement methods by plotting the differences against the averages of the measurements.

Because increasing levels of sleep disturbance have been shown to lower agreement between sleep devices and PSG,^31,32^ we evaluated the performance of the HB or ACT with PSG among subgroups of the poorest sleepers using the following thresholds: AHI ≥15, ISI ≥15, PSQI >5, and PLMI ≥15. Adherence and usability were assessed via descriptive statistics. All analyses were performed using SAS 9.4 (SAS Institute Inc., Cary, NC). All tests had statistical significance set at *p* < .05.

## RESULTS

The characteristics of the 63 participants who had complete data from the three sleep devices are shown in Table 1. The average age was 72.8 (standard deviation [SD] 6.6) years, 63.5% were female, and 63.5% identified as non-Hispanic White. Overall, participants had poor sleep, with average scores on the PSQI and ISI of 9.4 [3.9] and 12.7 [4.7], respectively. PSG-related sleep characteristics included averages for TST and WASO of 370.1 [93] and 88.9 [63] minutes, respectively, and an average AHI of 19.6 [15.3] events per hour.

**Table 1.** Demographic and clinical characteristics of the sample.

| Characteristics | N=63<br>Mean (SD) or n (%) |
| --- | --- |
| Age, years | 72.8 (6.6) |
| Female | 40 (63.5%) |
| Non-Hispanic White | 40 (63.5%) |
| College graduate | 25 (40.3%) |
| Retired | 44 (69.8%) |
| Body mass index | 28.35 (7.1) |
| Cognitive impairment <sup>a</sup> | 23 (36.5%) |
| History of diabetes <sup>b</sup> | 11 (17.5%) |
| History of coronary artery disease <sup>b</sup> | 7 (11.1%) |
| History of heart failure <sup>b</sup> | 5 (7.9%) |
| History of chronic obstructive pulmonary disease <sup>b</sup> | 9 (14.3%) |
| History of a psychiatric disorder <sup>c</sup> | 24 (38.1%) |
| History of sleep disorder <sup>b</sup> | 21 (33.3%) |
| Sleep Apnea <sup>d</sup> | 16 (25.4%) |
| Insomnia | 7 (11.1%) |
| Restless legs syndrome | 3 (4.8%) |
| Medication count <sup>e</sup> | 7.7 (4.8) |
| Reported use of a sleep aid in the past two weeks <sup>f</sup> | 19 (30.7%) |
| Pittsburgh Sleep Quality Index score | 9.4 (3.9) |
| Epworth Sleepiness Scale score | 7.9 (4.9) |
| Insomnia Severity Index score | 12.7 (4.7) |
| Total sleep time, min <sup>g</sup> | 370.1 (93.3) |
| Wake after sleep onset, min <sup>g</sup> | 88.9 (62.8) |
| Apnea hypopnea index, events/hour <sup>g</sup> | 19.6 (15.3) |
| Periodic limb movement index, events/hour <sup>g</sup> | 8.8 (16.9) |

<sup>a</sup> Score <12 on the Montreal Cognitive Assessment short form.
<sup>b</sup> Self-reported physician diagnosis
<sup>c</sup> Self-reported physician diagnosis of at least one of the following: depression (13), anxiety (17), bipolar disorder (1), post-traumatic stress disorder (3), substance use disorders (1).
<sup>d</sup> 2 participants used continuous positive airway pressure therapy during the study.
<sup>e</sup> Total number of medications taken in the past two weeks, including prescription medications and over-the-counter use of aspirin, pain medication (e.g., acetaminophen, ibuprofen, naproxen), allergy/cold medicines, or antacids (e.g., tums, famotidine, proton pump inhibitor); does not include use of bowel regimens, eyedrops, topical lotions, nasal sprays, multivitamins or probiotics.
<sup>f</sup> Inquired by asking, “In the past two weeks, have you taken a sleep aide to help you sleep during the night?” and includes prescription or over-the-counter medications or supplements.
<sup>g</sup> From polysomnography

Comparisons of sleep duration and quality from the HB or ACT with PSG are shown in Table 2. For the HB, the mean differences when compared to PSG for measurements of TST and WASO were −11.9 [−25.8, 2.0] minutes (*p*=0.09) and −15.5 [−29.5, −1.5] minutes ((*p*=0.03), respectively. ICCs for the HB compared with PSG showed good agreement for TST (0.78 [0.64, 0.85]) and moderate agreement for WASO (0.68 [0.48, 0.77]). Mean differences for ACT when compared to PSG were larger, with a mean difference for TST of −47.7 [−67.6, −27.8] minutes (*p* < .001) and a mean difference for WASO of 43.1 [27.9, 58.3] minutes (*p* < .001). ICCs for ACT compared with PSG showed moderate agreement for TST (0.52 [0.32, 0.68]) and poor agreement for WASO (0.13 [−0.1, 0.38]).

**Table 2.**
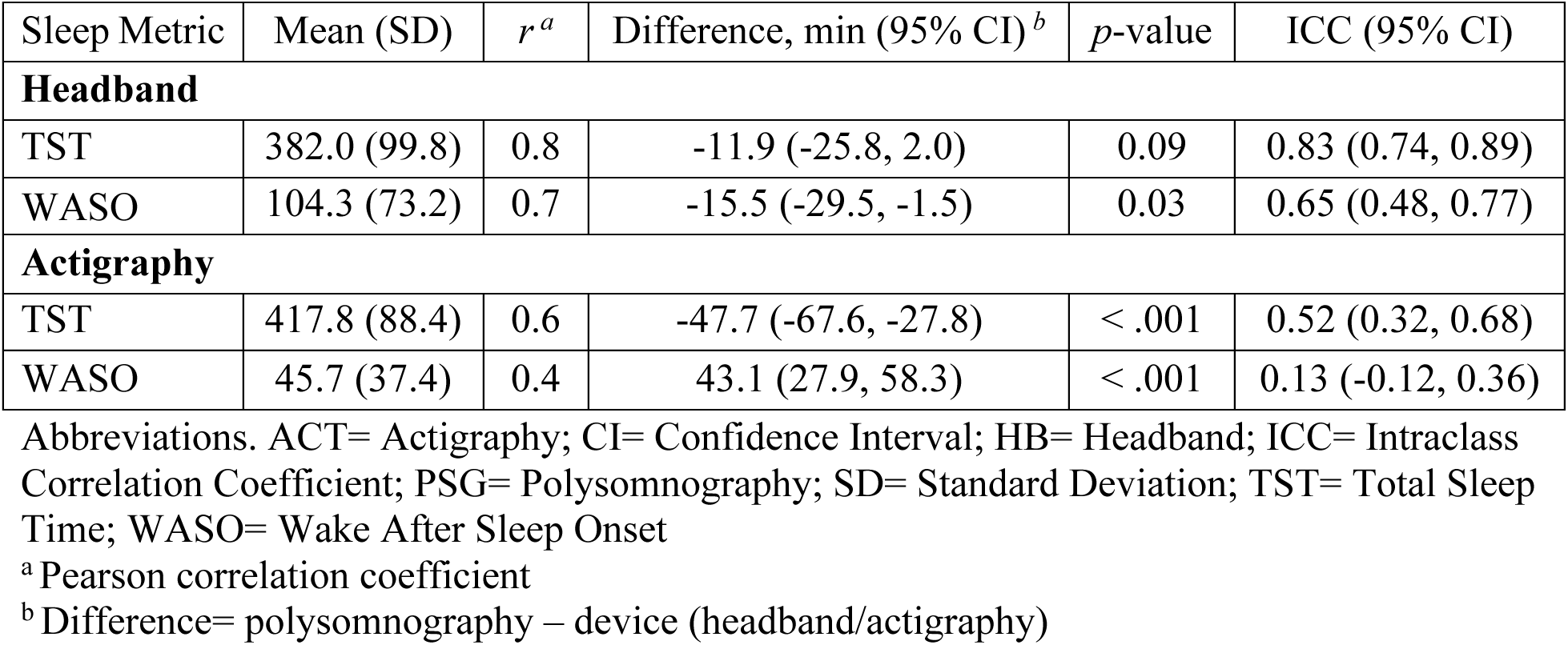
Comparisons of sleep duration and quality from the HB or ACT with PSG.

| Sleep Metric | Mean (SD) | $r^a$ | Difference, min (95% CI)<sup>b</sup> | $p$-value | ICC (95% CI) |
| --- | --- | --- | --- | --- | --- |
| **Headband** |  |  |  |  |  |
| TST | 382.0 (99.8) | 0.8 | -11.9 (-25.8, 2.0) | 0.09 | 0.83 (0.74, 0.89) |
| WASO | 104.3 (73.2) | 0.7 | -15.5 (-29.5, -1.5) | 0.03 | 0.65 (0.48, 0.77) |
| **Actigraphy** |  |  |  |  |  |
| TST | 417.8 (88.4) | 0.6 | -47.7 (-67.6, -27.8) | < .001 | 0.52 (0.32, 0.68) |
| WASO | 45.7 (37.4) | 0.4 | 43.1 (27.9, 58.3) | < .001 | 0.13 (-0.12, 0.36) |

Abbreviations. ACT= Actigraphy; CI= Confidence Interval; HB= Headband; ICC= Intraclass Correlation Coefficient; PSG= Polysomnography; SD= Standard Deviation; TST= Total Sleep Time; WASO= Wake After Sleep Onset
<sup>a</sup> Pearson correlation coefficient
<sup>b</sup> Difference= polysomnography – device (headband/actigraphy)

Bland-Altman plots displaying mean differences for measures of sleep duration and quality for the HB or ACT compared with PSG are shown in Figures 1 and 2. For the HB, plots in Figures 1a and 1b show a fixed systematic bias for over-estimating TST and WASO compared with PSG. For ACT, the plots for in Figures 2a and 2b show a larger systematic bias, with ACT over-estimating TST and under-estimating WASO compared with PSG. For Figure 2b, there is wider scatter at higher values when comparing measures of WASO between ACT and PSG, suggesting that differences between these devices increased as the magnitude of WASO increased. For all other plots, individual measures scattered randomly, suggesting that variability was uniform across the range of measurements. Both plots had wide 95% limits of agreement, with greater differences between ACT and PSG than between HB and PSG.

**Figure 1.**
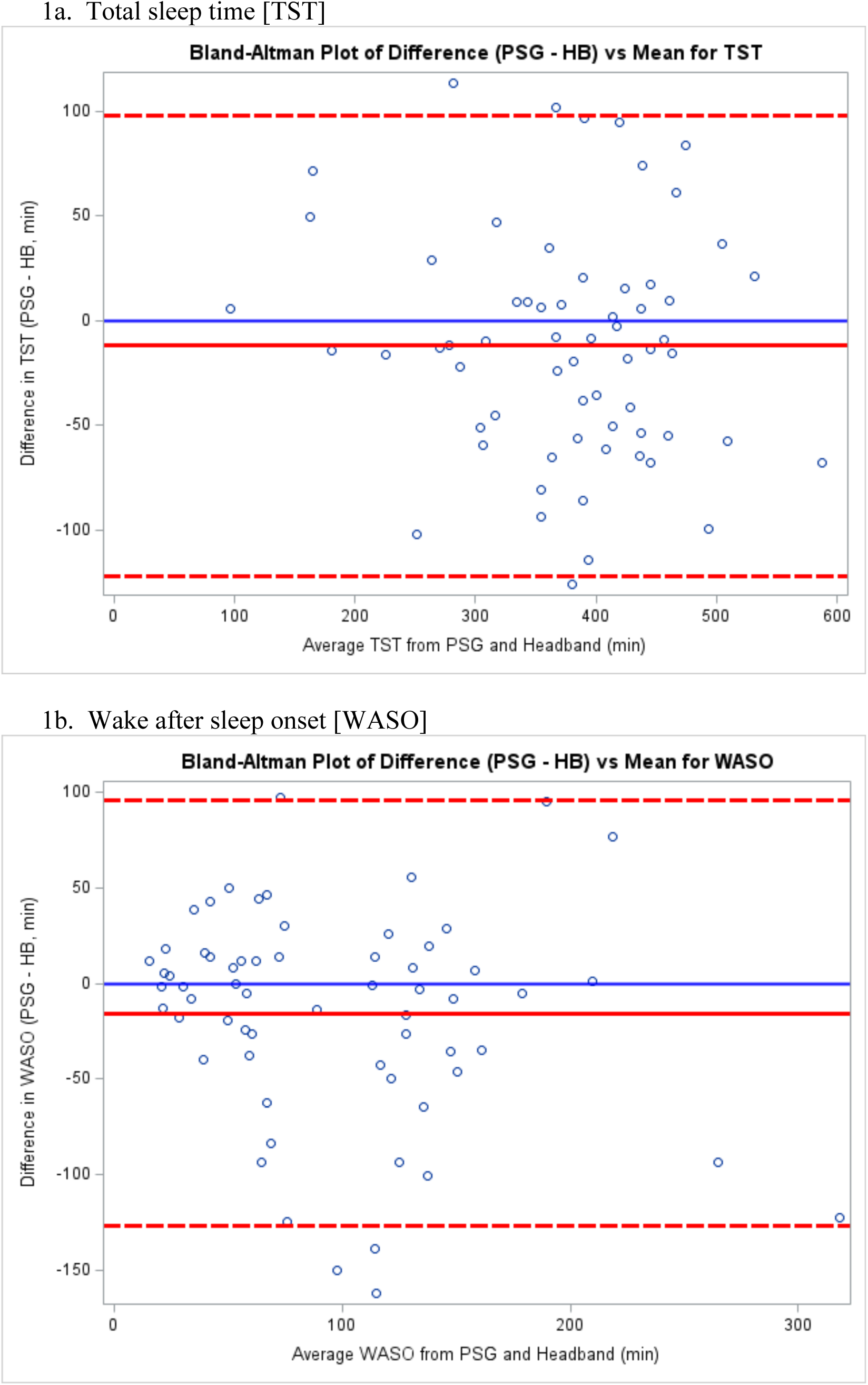
Bland-Altman plots comparing measures of sleep duration and quality from the HB with PSG. Plots show the agreement of the HB compared with PSG for TST (1a) and WASO (1b). The x-axis represents the average of TST or WASO from the two devices, while the y-axis represents the difference between the two devices (i.e., PSG - HB). The solid line represents the mean difference, while the dashed lines indicate the limits of agreement (mean difference ± 1.96 SD). Each point represents a measurement pair.

**Figure 2.**
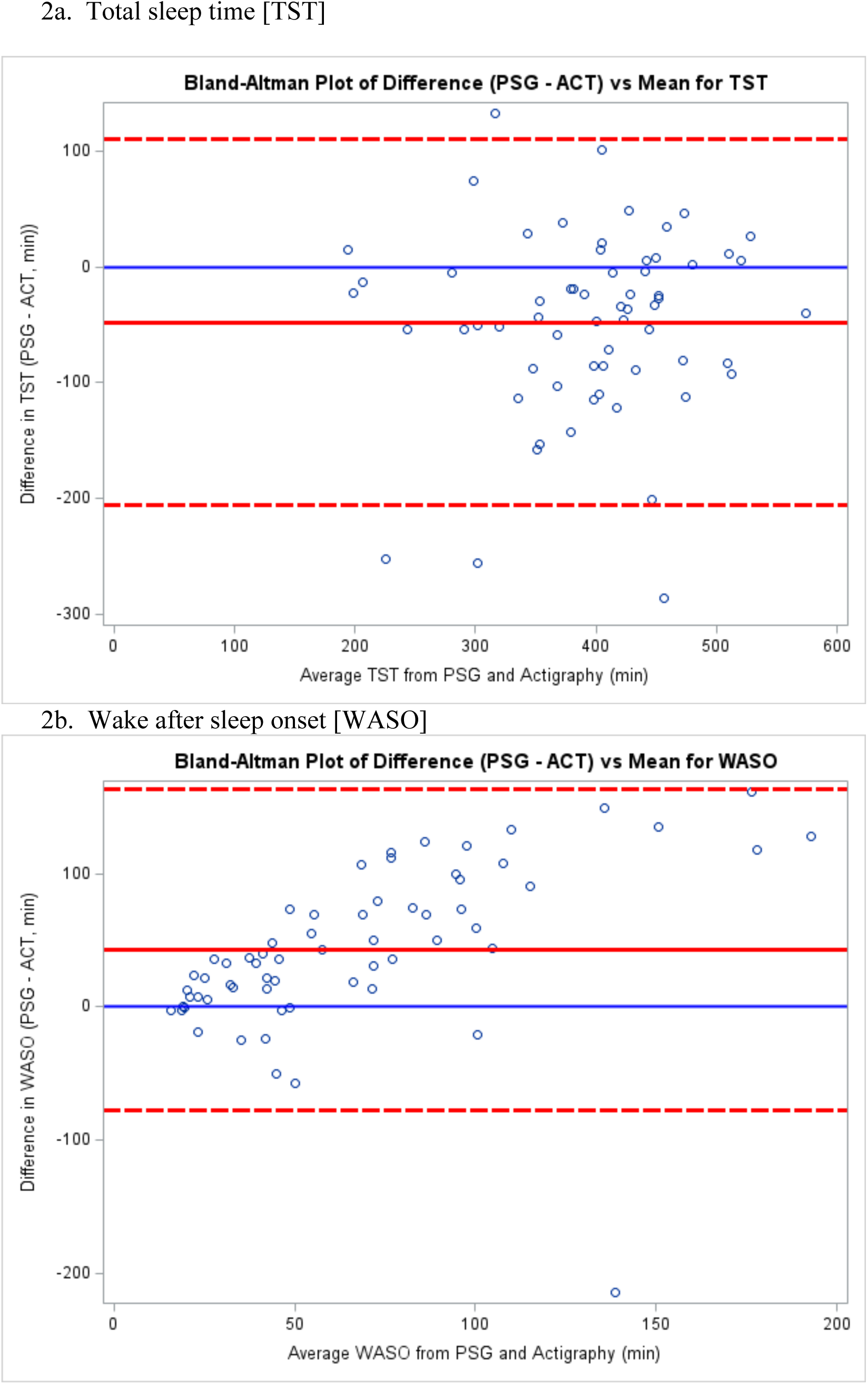
Bland-Altman plots comparing measures of sleep duration and quality from ACT with PSG. Plots show the agreement of the ACT compared with PSG for TST (2a) and WASO (2b). The x-axis represents the average of TST or WASO from the two devices, while the y-axis represents the difference between the two devices (i.e., PSG - ACT). The solid line represents the mean difference, while the dashed lines indicate the limits of agreement (mean difference ± 1.96 SD). Each point represents a measurement pair. Abbreviations: ACT=Actigraph; HB= Headband; PSG=Polysomnography; TST=total sleep time; WASO=wake after sleep onset.

Table 3 shows the performance of the HB or ACT compared with PSG among the poorest sleepers, defined according to well-accepted thresholds from validated questionnaires or PSG. Among persons with sleep apnea (AHI ≥15), moderate to severe insomnia (ISI ≥15), poor sleep quality (PSQI>5), or increased leg movements during sleep (PLMI ≥15), the HB showed good to excellent agreement with PSG for TST and moderate to good agreement for WASO. In contrast, ACT showed lower levels of agreement with PSG among the poorest sleepers, with ICCs ranging 0.49 to 0.52 for TST and all ICCs <0.20 for WASO.

**Table 3.**
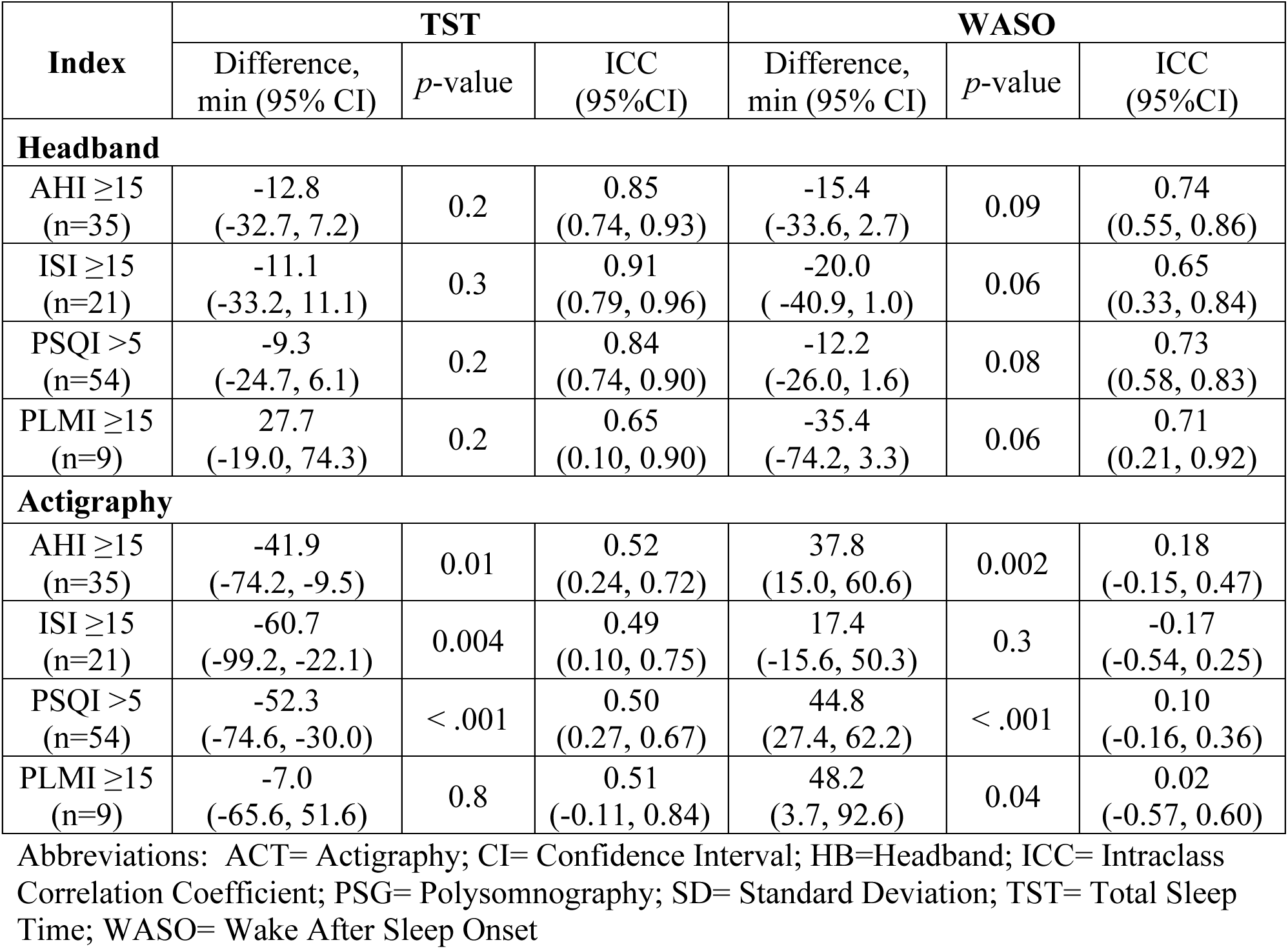
Agreement of headband- or actigraphy-assessed duration and quality with PSG among the poorest sleepers.

Adherence and acceptability data for the HB are shown in Table 4. On average, participants wore the HB for 6.5 [0.8] nights. Fifty-nine of the participants (93.7%) wore the HB for ≥four nights and 39 participants (61.9%) wore the HB for seven nights. Median scores for usability for each WEAR scale item were 4 and above (i.e., agree or strongly agree).

**Table 4.** Adherence and Usability Data for the HB.

| **Adherence** | **Mean (SD)**<br>**or n (%)** |
| --- | --- |
| Total Nights Worn | 6.5 (0.8) |
| 4+ Hours/Night Adherence | 59 (93.7) |
| 7 Nights Adherence <sup>a</sup> | 39 (61.9) |
| **Usability <sup>b</sup>** | **Median**<br>**(IQR)** |
| 1. The sleep headband seems to be useful and easy to use. | 4 (3, 5) |
| 2. The sleep headband seems like "too much" technology.* | 5 (4, 5) |
| 3. The sleep headband restricts movement or physically gets in the way.* | 4 (3, 5) |
| 4. I needed the support of a technical person to be able to use the headband.* | 5 (4, 5) |
| 5. The sleep headband seems comfortable, not bulky. | 4 (3, 5) |
| 6. The sleep headband is sleek, not clunky. | 4 (3, 4) |
| 7. I would imagine that most people would learn to use the sleep headband very quickly. | 4 (3, 5) |
| 8. I found the sleep headband very cumbersome to use.* | 4 (3, 5) |
| 9. I felt very confident using the sleep headband. | 4 (3, 5) |
| 10. I needed to learn a lot of things before I could get going with the sleep headband.* | 5 (4, 5) |
| 11. I think the sleep headband could help people. | 4 (4, 5) |

Abbreviations: HB=Headband; SD=standard deviation
<sup>a</sup> 63 (98.4%) participants wore the headband for at least 4 nights
<sup>b</sup> From the WEearable Acceptability Range (WEAR) scale

## DISCUSSION

Among 63 community-dwelling older adults with symptoms of insomnia and/or daytime sleepiness, we found that an EEG-HB out-performed ACT, providing measurements of sleep that were more consistent with PSG. This was true even among the poorest sleepers, including persons with sleep apnea or moderate-to-severe insomnia, in whom accurate measures are essential and for whom ACT may be less reliable. Our study population was able to wear the HB over multiple nights and rated its usability highly. These results support the use of the HB or similar EEG-based devices to provide measures of sleep with increased accuracy in this population.

This study contributes to previous literature validating the HB, albeit in different populations. In an initial study by Arnal *et al.* among 25 healthy, middle-aged adults, the HB showed an overall sleep staging accuracy of 83.8% compared to consensus sleep staging from PSG.^33^ Another study by Gonzalez *et al.* among a clinical population of 10 persons with Parkinson’s disease found 80.8% accuracy across all stages for the HB compared to PSG.^34^ As in our study, Gonzalez *et al.* found that the HB overestimated TST and showed, in an epoch-by-epoch analysis, that the HB significantly overestimated time in deep sleep and rapid eye movement (REM) sleep compared to PSG. Gonzalez *et al.* concluded that the HB had high specificity but lower sensitivity for identifying specific sleep stages.^34^ A third study by Wood *et al.* compared a newer version of the HB, the “Dreem 3”, to another consumer device and to sleep diaries among 25 college students.^35^ While the authors did not have PSG data, they used a complex analysis based on previously reported data from Arnal *et al.* to estimate a lower limit for the expected epoch-by-epoch sleep stage agreement of 78.1% vs. PSG. The authors also reported that the HB tended to underestimate periods of wakefulness and mistake it for light sleep (i.e., non-REM stages 1 and 2).^35^ While the results of the studies from Gonzalez and Wood should be interpreted with caution in light of small sample sizes and high attrition rates, their results and ours suggest that the HB performs well compared to PSG but may overestimate TST due to difficulties distinguishing wake from lighter sleep stages, and may also overestimate time in deep and REM sleep. Our study extends findings by demonstrating the HB’s robust performance in a larger and more diverse and generalizable sample of older adults with sleep disturbances.

Our study is the first to simultaneously compare the performance of the HB and ACT, which is the current reference standard for assessment of sleep over multiple nights. Known limitations of ACT include its bias towards overestimation of TST and underestimation of WASO in all persons, as well as its tendency to perform worse among persons with multiple chronic conditions and sleep disturbances.^16,31,32^ ACT relies on accelerometry to record movements and infers sleep periods via specialized algorithms. Thus, it is prone to errors of overestimation of TST in persons who may be awake but trying to sleep (e.g., persons with insomnia) but also to underestimation of TST among persons who may move more during sleep (e.g., persons with sleep apnea or leg movements during sleep). Conversely, the HB’s ability to measure EEG activity assesses sleep directly and may explain its superior performance, especially among the poorest sleepers. Thus, the HB may be more appropriate for studies requiring accurate sleep metrics among persons with disturbed sleep, while ACT may be better used as a tool to provide objective, longitudinal patterns of sleep in persons without major sleep disturbances or when 24-hour rest-activity patterns are the study focus.

The feasibility of the HB has been assessed previously, although in younger populations. In a study of the feasibility of the HB among 21 adults with chronic pain (average age 44 years), 86% of participants said they would wear the HB longer than the 2-night minimum requirement, and over half of the sample rated the HB as comfortable while sleeping.^36^ While some might assume that an older population might have more difficulty adapting to new technologies, we show high rates of feasibility and acceptability in this group. Importantly, our inclusion criteria required that participants identified as having a sleep problem, a population in whom successful use of a wearable device to assess sleep should not be assumed. Nevertheless, most of our participants could sleep with the HB, as evidenced by 95.2% of participants wearing the HB for ≥4 nights. However, two issues are important to note. The first is that among the 71 enrolled participants, six (8.5%) dropped out of the study due to inability to use the HB, citing difficulty turning on the HB or inability to sleep with the HB as the reason for dropping out. Thus, accounting for this group of early drop-outs provides a better expectation of feasibility. Second, we did not require participants to use a mobile device to manage the HB. Doing so would have limited our ability to study a more generalized population and may have led to more difficulties with HB use. It is important to note that the manual start for recordings on the newer generation of headbands is easier and that individuals who are comfortable with smartphone technology may find that the mobile devices increases feasibility by making it easier to begin and end recordings.

While the HB shows promising results with respect to measures of sleep duration, additional work is needed to improve its measurement of sleep fragmentation (e.g., WASO) and of sleep stages. Because raw EEG data is available from the HB, one might consider direct scoring of sleep data by a trained sleep technologist rather than relying on the HB’s autoscoring algorithm. However, a preferred approach may be to improve the algorithm, as this would increase feasibility and scalability in future studies. Thus, an essential next step of our work will be to complete an epoch-by-epoch analysis for the sleep staging of the HB compared with PSG, which will contribute to improved performance of the algorithm. Additional steps to improve the algorithm may also include optimizing training data through additional studies that provide high-quality, well-labeled data from diverse sources; refining preprocessing of data through signal filtering and artifact removal; incorporation of multimodal data (e.g., from EOG or EMG) to improve accuracy; and continued external validation from different populations and environments. The HB’s autoscoring algorithm and the ability to provide data on sleep staging are major advantages compared to other wearable devices, and improvements to its performance have the potential to lead to significant advances in the field of sleep medicine.

This study’s strengths include its focus on a generalizable sample of community-living older adults, high representation of minorities, high retention rates, and simultaneous comparison of multiple sleep devices on the same night in the home environment. Furthermore, the inclusion of participants with known sleep disorders and use of hypnotic medications enhances the validity of our findings. However, several limitations are important to note. First, the sample size, while larger than in previous studies, is modest and predominantly composed of highly educated individuals. Second, our analysis focused on TST and WASO. A detailed epoch-by-epoch analysis is needed to enhance our understanding of the sensitivity and specificity of the HB’s sleep staging compared with PSG. Third, our usability should be interpreted with caution. As noted above, several participants dropped out of the study due to difficulties using the HB. In addition, we adapted our protocol for the study population (e.g., not requiring participants to use a mobile device). Others may find that mobile devices increase or decrease feasibility depending on the population being studied. Finally, we studied the “Dreem 2” device, which has been replaced by the “Dreem 3”; thus, our performance and usability data may not be directly applicable to the newer device.

In summary, we found that an EEG-measuring HB demonstrated good agreement with PSG for measures of sleep duration, outperforming ACT, especially among the poorest sleepers. Additional validation of the HB is needed to improve the identification of WASO and specific sleep stages. Devices like the HB might provide more accurate and feasible measures of sleep in habitual sleep environments, enhancing the evaluation and management of sleep health in older adults.

## Data Availability

All data produced in the present study are available upon reasonable request to the authors.

## ACKNOWLEDGEMENTS

No funding was provided by the manufacturer of the devices tested in this study. Additionally, we declare that there are no conflicts of interest related to this work.

